# Mental sequelae of the Covid-19 pandemic: Well-being one year into the crisis in children with and without complex medical histories and their parents

**DOI:** 10.1101/2021.12.05.21267236

**Authors:** Melanie Ehrler, Cornelia F. Hagmann, Oliver Kretschmar, Markus A. Landolt, Beatrice Latal, Flavia M. Wehrle

## Abstract

**Objectives:** To understand the long-term mental sequelae for families over the course of the COVID-19 pandemic, the well-being of children with and without complex medical histories and their parents was investigated longitudinally.

**Methods:** Well-being of 200 school-aged children (73 typically-developing, 46 born very preterm, 73 with complex congenital heart disease) and 175 of their parents was assessed prior to and during the first (April–May 2020), second (October–November 2020), and third waves (April–May 2021) of the pandemic with standardized questionnaires. Social and COVID-19-specific determinants were investigated as predictors of impaired well-being.

**Results:** Child proxy-reported well-being was lower than before the pandemic during the first (*P*<0.001) and third waves (*P*=0.01) but not the second (*P*=0.13). Child self-reported well-being was not lower during the pandemic (all *P*>0.10). Parent well-being dropped during the first wave (*P*<0.001) and remained low during the first year (*P*<0.01). One year into the pandemic, 18/25% of children (self-/proxy-report) and 27% of parents scored below the normal range compared to 11%/10%, and 16%, respectively, before the pandemic. Parents of typically-developing children reported lower well-being than parents of children born very preterm (*P*=0.006) or with a complex congenital heart disease (*P*=0.03). Child and parent well-being was lower in families with sparse social support (*P*<0.001) and poor family functioning (*P*<0.01).

**Conclusion:** The pandemic continues to impact family well-being one year after its outbreak. Families with sparse social support and poor family functioning are particularly at risk for compromised well-being and support should be provided to them.

## 1. Introduction

More than a year into the COVID-19 pandemic, evidence for an acute negative impact on mental health has accumulated around the globe (see e.g., [1, 2] for an overview). From the beginning of this crisis, children and adolescents were identified as being particularly at risk for impaired well-being due to the profound changes in the psychosocial environment that accompanied measures to halt the spread of the pandemic, particularly the closing of schools and the reduction of social contacts [3-7]. Indeed, numerous studies have reported reduced well-being and high rates of internalizing and externalizing problems and symptoms of anxiety and depression in children and adolescents during the first wave [8-20]. Parents were also strongly burdened by the pandemic: many of them faced increased parental responsibilities and stress working remotely while concurrently caring for their children at home [15, 19, 21-27]. Indeed, the well-being of parents was reported to be more strongly affected than that of adults without children [28, 29]. A number of factors have been associated with poor well-being during the pandemic, including social determinants such as low socio-economic status (SES) and sparse social support [1]. In addition, it is unclear whether children with pre-existing medical conditions and their parents are at particular risk for lower well-being [13, 19, 30].

To date, the overwhelming majority of studies reported on the acute impact of the COVID-19 pandemic on well-being during the initial wave in early 2020. However, the long-term mental sequelae of this ongoing pandemic remain unknown. Thus, the aim of the current study was to investigate the well-being of children and their parents as the pandemic evolved. Data from two cohort studies assessed prior to the outbreak was complemented with data collected at three time-points over the course of the pandemic to investigate the immediate (first wave, April–May 2020), intermediate (second wave, October–November 2020), and long-term (third wave, April–May 2021) impact on children and parents and to identify factors contributing to impaired well-being.

## 2. Methods

### 2.1. Participants and study procedure

Families were recruited from two prospective cohort studies at the University Children’s Hospital Zurich, Switzerland. The aim of those studies is to assess the neurodevelopmental outcome of children at school age who are either typically-developing or born very preterm (VPT) or who have a complex congenital heart disease [31, 32]. Families were eligible for this current study if the child had participated in the neurodevelopmental assessment and the parents had completed a set of questionnaires on child and parent well-being between January 2013 and mid-March 2020 as part of one of the cohort studies (T0: prior to the implementation of measures to reduce COVID-19 in Switzerland). Parents of eligible families were invited to complete an online survey once during the first wave of the pandemic (T1: between April 17 and May 10, 2020), while lockdown measures, including school closure, were in place in Switzerland[13]. Families who had participated in the T1 assessment were approached again once during the second wave (T2: between October 30 and November 22, 2020) and once during the third wave (T3: between April 23 and May 23, 2021), when governmental restrictions were less severe: schools were open, but public and private assemblies were restricted. Supplementary Fig1 details the assessment procedure for the current study. Supplementary Table 1 lists the restriction measures in Switzerland at each assessment time-point.

Families who participated in the COVID-19 survey at T1 did not differ in family SES or parent well-being prior to the pandemic (T0) from those who did not [13]. Further, parents who participated at T2 (*P*=0.192) and T3 (*P*=0.671) did not differ in parent well-being at T1 from those who did not participate. The study was approved by the local ethics committee, and all parents gave written informed consent.

### 2.2. Assessment instruments

Standardized questionnaires assessing quality of life were selected from the protocols of the two cohort studies (T0) and were included in the online surveys at T1, T2, and T3.

For children, the *psychological well-being* subscale of *Kidscreen-27 [33]* was used to assess well-being. Parents completed the proxy-report of the scale at all time-points. Children completed the self-report of the scale at T0, T2 and T3. At T0, children completed the questionnaires during the on-site assessment for the prospective cohort studies. At T2 and T3, the questionnaires were sent to the children by mail after the parents had completed the online survey. The *psychological well-being* subscale includes 7 items that assess the child’s positive emotions and satisfaction and the absence of feelings of loneliness and sadness. Raw subscale scores were transformed into T-values with Swiss norms (n = 1672, adjusted for age and sex). Low values indicate poor well-being [33]. For parents, the *mental* subscale of the *Short Form Health* questionnaire was used to assess self-reported well-being. The 36-item version (*SF-36*) was used at T0, and the 12-item short form (*SF-12*) was used at T1, T2, and T3 [34]. For the analysis at T0, only the 12 items overlapping with the short form were considered. The *mental* subscale of the *SF-12* assesses four dimensions: *vitality, social function, role limitations due to emotional problems*, and *mental health*. Raw scores were transformed into T-values based on German norms (n = 2524) [35]. Both the *Kidscreen-27* and the *SF-12* have acceptable to good internal consistency [33, 34].

Several potential predictors of child and parent well-being were assessed: Family SES was defined as a combined score of maternal and paternal education. Higher scores indicate higher SES (range 2 to 12). At T0, parents completed the 14-item short form of the *Social Support Questionnaire* (*F-SozU K14* [36]), assessing the perceived extent of support from the social network that is accessible if needed. Three dimensions, *emotional support, practical support*, and *social integration*, were summed according to the manual. Higher scores indicate more social support. The *F-SozU K14* has excellent internal consistency [36]. The quality of family functioning was assessed during the first wave of the pandemic with the 27-item *Family Relationship Index* (*FRI* [37]). Three dimensions, *cohesion, expressiveness*, and *conflict*, were summed according to the manual. Higher scores indicate better quality of family functioning. The FRI has good internal consistency [37]. Familial COVID-19 risk status was assessed by asking parents whether a family member was at risk for a severe disease course in case of an infection with SARS-Cov-2 due to a pre-existing health condition. A dichotomous variable differentiated families with a member at increased risk from those without.

### 2.3. Statistical Analysis

Parent and child characteristics are expressed as numbers and proportions of totals (dichotomous data), median and interquartile range (ordinal data), and mean and standard deviation (continuous data). Child and parent well-being were compared to normative data at each time-point using one-sample *t*-tests for normally distributed data and Mann–Whitney *U-*tests for skewed data. Effect sizes were estimated with Cohen’s *d* (small effect >0.2, moderate effect >0.5, strong effect >0.8 [38]). The proportion of children and parents scoring below the normal range (>1 SD below the normative mean or median) were reported separately for each time-point.

Longitudinal changes of child and parent well-being were investigated with mixed-effect models: Three models were calculated with the following outcomes: 1) proxy-reported child well-being, 2) self-reported child well-being, and 3) parent well-being. As fixed effects, the models included assessment time (categorical: T0 = prior to the pandemic; T1 = first wave, spring 2020; T2 = second wave, fall 2020; T3 = third wave, spring 2021), group (categorical: typically-developing, CHD, and VPT), and age at assessment of the child or the parent. As random effect, family-specific intercepts were included to take respondent-specific variability and shared variance between siblings into account (pairs of siblings: *n=*21). Children’s individual intercepts were nested within families. The potential predictors of child and parent well-being were then added to those models with a significant time effect.

Unstandardized regression coefficients (*B*) and generalized semi-partial *R*^2^ (*R*^*2*^_*B*_), to quantify effect sizes for mixed models, were reported (small: *R*^*2*^_*B*_<0.01; medium: *R*^*2*^_*B*_>0.09; large: *R*^*2*^_*B*_>0.25 [39]). The distribution of residuals was examined to evaluate normality.

All analyses were performed with *R* version 4.0.3 [40]. *P*-values at an α-level of 0.05 were considered statistically significant.

## 3. Results

### 3.1. Sample characteristics

Before the pandemic (T0), families of 346 children had participated in one of the two cohort studies and thus were eligible for the current study. Families of 200 children participated in the online survey at T1 (follow-up rate = 58%). At T2, families of 138 children (follow-up rate = 70%) completed the online survey again and at T3, families of 134 children (follow-up rate = 67%). Primarily, mothers reported on the well-being of their children (>90%). The questionnaires of both parents were available for 27 children, therefore only the mothers’ responses were retained for further analyses. In a number of families, parents completed the survey for more than one child, resulting in 175, 122, and 117 parents participating at T1, T2, and T3. Sample characteristics are displayed in Table 1.

**Table 1:**
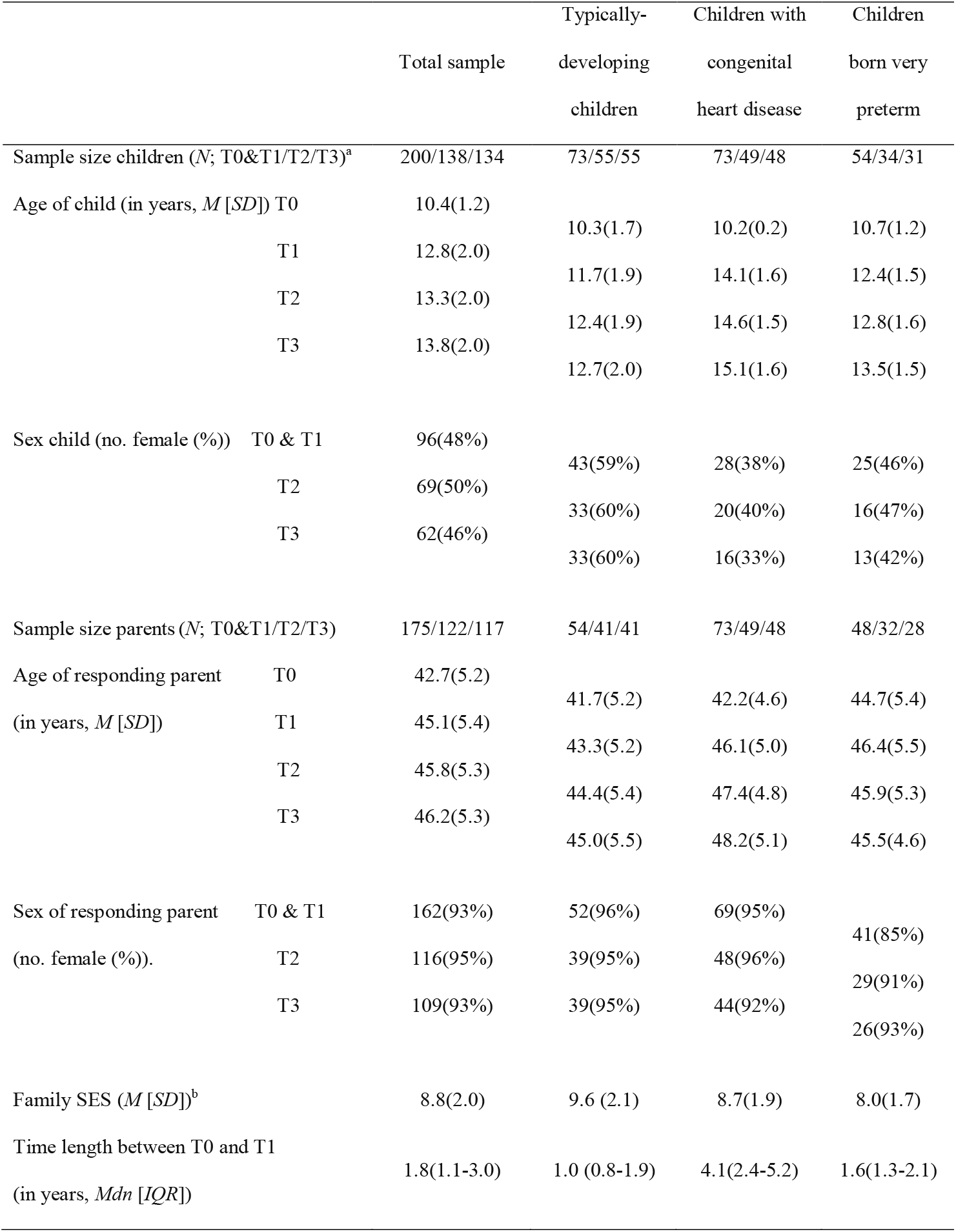
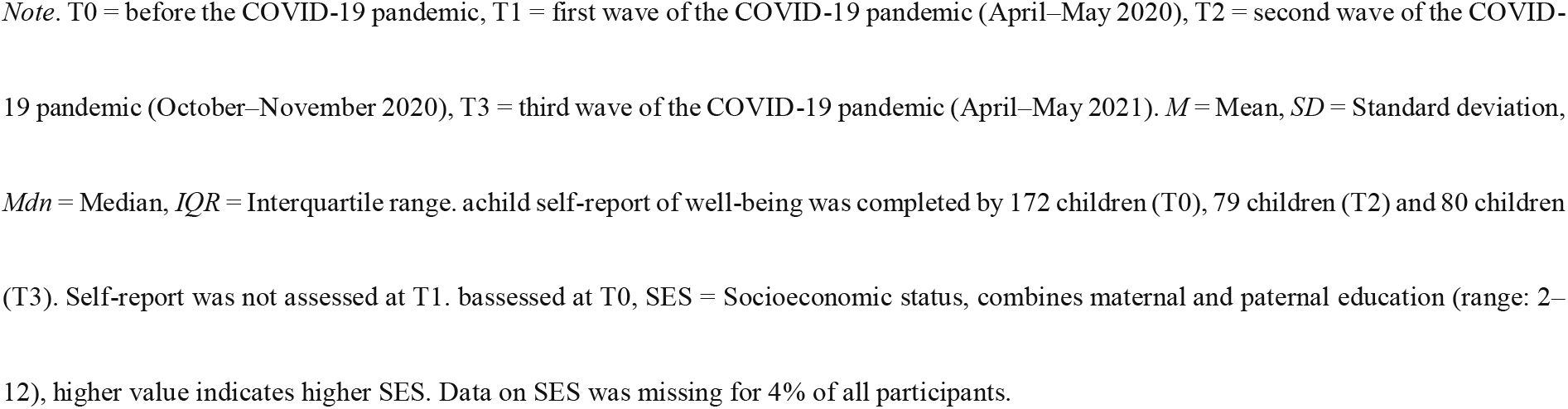
Sample characteristics

Of the children with CHD, 21% had a univentricular heart defect. Children born VPT were born at a mean gestational age of 28.9 weeks (SD=1.6). In 23 families, at least one family member had been infected with SARS-CoV-2. In 75 families, at least one family member was reported to be at risk for a severe course of disease in case of a SARS-CoV-2 infection.

### 3.2. Well-being of children and their parents before and during the COVID-19 pandemic

Table 2 displays the comparison to normative data for child and parent psychological well-being. Statistical estimates of the three models assessing longitudinal changes in child and parent well-being are presented in Table 3. Compared to before the pandemic, child proxy-reported well-being was significantly lower during the first and the third but not the second wave of the pandemic (Fig1A). The model’s effect size was small (*R*^*2*^_*B*_(CI-95)*=*0.047(0.027 to 0.090)). Adding the predictors increased the models effect size to medium (*R*^*2*^_*B*_(CI-95)*=*0.145(0.110 to 0.205)). Well-being was independent of group and age of the child. Sparse social support before the pandemic and poor family functioning during the first wave significantly predicted lower well-being. There was no significant interaction between either assessment time and perceived social support (*P*=0.084) or assessment time and family functioning (*P*=0.644) on well-being.

**Table 2:**
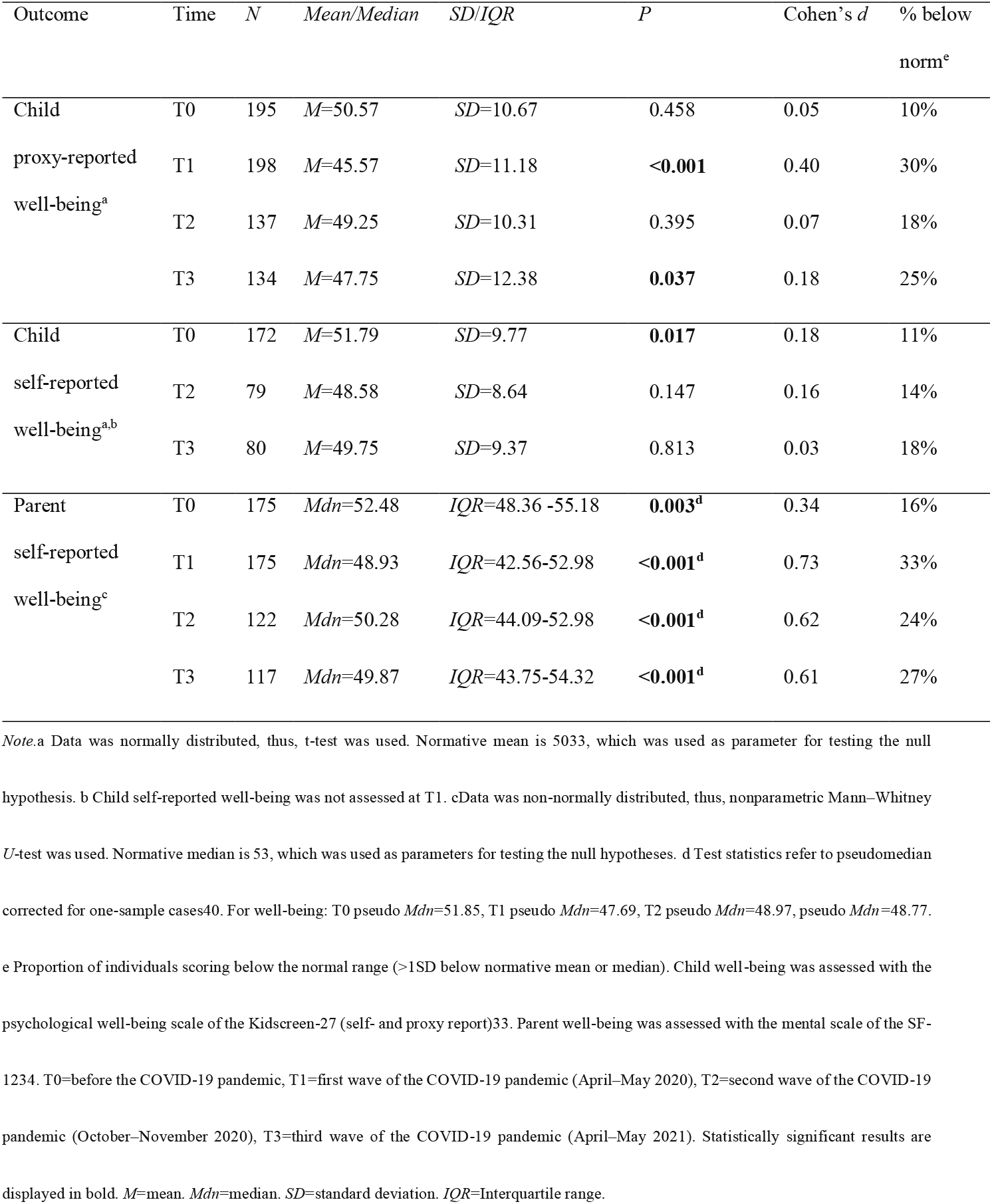
Well-being compared to normative data.

| Outcome | Time | *N* | *Mean/Median* | *SD/IQR* | *P* | Cohen's *d* | % below<br>norm<sup>e</sup> |
| --- | --- | --- | --- | --- | --- | --- | --- |
| Child | T0 | 195 | *M*=50.57 | *SD*=10.67 | 0.458 | 0.05 | 10% |
| proxy-reported | T1 | 198 | *M*=45.57 | *SD*=11.18 | **<0.001** | 0.40 | 30% |
| well-being<sup>a</sup> | T2 | 137 | *M*=49.25 | *SD*=10.31 | 0.395 | 0.07 | 18% |
|  | T3 | 134 | *M*=47.75 | *SD*=12.38 | **0.037** | 0.18 | 25% |
| Child | T0 | 172 | *M*=51.79 | *SD*=9.77 | **0.017** | 0.18 | 11% |
| self-reported | T2 | 79 | *M*=48.58 | *SD*=8.64 | 0.147 | 0.16 | 14% |
| well-being<sup>a,b</sup> | T3 | 80 | *M*=49.75 | *SD*=9.37 | 0.813 | 0.03 | 18% |
| Parent | T0 | 175 | *Mdn*=52.48 | *IQR*=48.36 -55.18 | **0.003<sup>d</sup>** | 0.34 | 16% |
| self-reported | T1 | 175 | *Mdn*=48.93 | *IQR*=42.56-52.98 | **<0.001<sup>d</sup>** | 0.73 | 33% |
| well-being<sup>c</sup> | T2 | 122 | *Mdn*=50.28 | *IQR*=44.09-52.98 | **<0.001<sup>d</sup>** | 0.62 | 24% |
|  | T3 | 117 | *Mdn*=49.87 | *IQR*=43.75-54.32 | **<0.001<sup>d</sup>** | 0.61 | 27% |

*Note.*a Data was normally distributed, thus, t-test was used. Normative mean is 5033, which was used as parameter for testing the null hypothesis. b Child self-reported well-being was not assessed at T1. cData was non-normally distributed, thus, nonparametric Mann–Whitney *U*-test was used. Normative median is 53, which was used as parameters for testing the null hypotheses. d Test statistics refer to pseudomedian corrected for one-sample cases<sup>40</sup>. For well-being: T0 pseudo *Mdn*=51.85, T1 pseudo *Mdn*=47.69, T2 pseudo *Mdn*=48.97, pseudo *Mdn*=48.77. e Proportion of individuals scoring below the normal range (>1SD below normative mean or median). Child well-being was assessed with the psychological well-being scale of the Kidscreen-27 (self- and proxy report)<sup>33</sup>. Parent well-being was assessed with the mental scale of the SF-1234. T0=before the COVID-19 pandemic, T1=first wave of the COVID-19 pandemic (April–May 2020), T2=second wave of the COVID-19 pandemic (October–November 2020), T3=third wave of the COVID-19 pandemic (April–May 2021). Statistically significant results are displayed in bold. *M*=mean. *Mdn*=median. *SD*=standard deviation. *IQR*=Interquartile range.

**Table 3:** Statistical estimates of fixed effects

| Variables | *B* | *CI-95* | *P* | *R*<sup>2</sup>*B*<sup>°</sup> | *B* | *CI-95* | *P* | *R*<sup>2</sup>*B*<sup>°</sup> |
| --- | --- | --- | --- | --- | --- | --- | --- | --- |
|  | *Simple model* |  |  |  | *Model including predictors* |  |  |  |
| Child proxy-reported well-being |  |  |  |  |  |  |  |  |
| Time <sup>a</sup> |  |  |  |  |  |  |  |  |
| T1 | -4.88 | -6.91,-2.85 | <0.001 | 0.028 | -5.33 | -7.66,-3.00 | <0.001 | 0.027 |
| T2 | -1.30 | -3.68, 1.09 | 0.288 | 0.002 | -2.10 | -4.81, 0.61 | 0.133 | 0.003 |
| T3 | -2.83 | -5.40,-0.27 | 0.031 | 0.007 | -3.92 | -6.87,-0.96 | 0.010 | 0.011 |
| Age | -0.01 | -0.55, 0.53 | 0.972 | 0.000 | 0.23 | -0.40, 0.86 | 0.479 | 0.001 |
| Group <sup>b</sup> |  |  |  |  |  |  |  |  |
| VPT born | 2.16 | -1.11, 5.42 | 0.198 | 0.007 | 3.25 | -0.20, 6.71 | 0.068 | 0.012 |
| CHD | 1.78 | -1.43, 4.99 | 0.278 | 0.005 | 0.71 | -2.62, 4.05 | 0.676 | 0.001 |
| SES at T0 |  |  |  |  | 0.30 | -0.41, 1.01 | 0.406 | 0.003 |
| Perceived social support at T0 |  |  |  |  | 0.09 | 0.04, 0.14 | <0.001 | 0.041 |
| Family functioning at T1 |  |  |  |  | 0.55 | 0.21, 0.90 | <0.001 | 0.037 |
| COVID risk status of family |  |  |  |  | 1.60 | -1.11, 4.30 | 0.252 | 0.005 |
| Child self-reported well-being <sup>c</sup> |  |  |  |  |  |  |  |  |
| Time <sup>a,d</sup> |  |  |  |  |  |  |  |  |
| T2 | -2.53 | -5.50, 0.45 | 0.098 | 0.005 |  |  |  |  |
| T3 | -1.27 | -4.46, 1.93 | 0.439 | 0.001 |  |  |  |  |
| Age | -0.19 | -0.86, 0.48 | 0.579 | 0.001 |  |  |  |  |
| Group <sup>b</sup> |  |  |  |  |  |  |  |  |
| VPT born | 2.12 | -0.75, 4.98 | 0.150 | 0.004 |  |  |  |  |
| CHD | 1.57 | -1.00, 4.14 | 0.233 | 0.003 |  |  |  |  |
| Parent well-being |  |  |  |  |  |  |  |  |
| Time <sup>a</sup> |  |  |  |  |  |  |  |  |
| T1 | -3.67 | -5.16,-2.19 | <0.001 | 0.022 | -3.48 | -5.02,-1.94 | <0.001 | 0.018 |
| T2 | -2.58 | -4.27,-0.89 | 0.003 | 0.016 | -2.60 | -4.32,-0.89 | 0.003 | 0.009 |
| T3 | -3.54 | -5.30,-1.79 | <0.001 | 0.009 | -3.58 | -5.36,-1.80 | <0.001 | 0.016 |
| Age | -0.02 | -0.22, 0.18 | 0.827 | 0.000 | 0.04 | -0.16, 0.24 | 0.713 | 0.000 |

| Group<sup>b</sup> |  |  |  |  |  |  |  |  |
| --- | --- | --- | --- | --- | --- | --- | --- | --- |
| VPT born | 3.44 | 0.76, 6.12 | **0.013** | 0.020 | 4.13 | 1.23, 7.03 | **0.006** | 0.023 |
| CHD | 1.96 | -0.55, 4.47 | 0.127 | 0.008 | 2.91 | 0.25, 5.57 | **0.034** | 0.014 |
| SES at T0 |  |  |  |  | 0.02 | -0.57, 0.61 | 0.950 | 0.000 |
| Perceived social support at T0 |  |  |  |  | 0.07 | 0.03, 0.12 | **<0.001** | 0.038 |
| Family functioning at T1 |  |  |  |  | 0.42 | 0.14, 0.70 | **0.004** | 0.027 |
| COVID risk status of family |  |  |  |  | -1.16 | -3.39, 1.07 | 0.310 | 0.003 |

Note. T0 = before the COVID-19 pandemic, T1 = first wave of the COVID-19 pandemic (April–May 2020), T2 = second wave of the COVID-
19 pandemic (October–November 2020), T3 = third wave of the COVID-19 pandemic (April–May 2021). Statistically significant results are
displayed in bold. *B* = unstandardized estimate. *CI-95* = 95% confidence interval. *P* = *P* value. VPT = very preterm born. CHD = congenital
heart disease. a Reference = T0. b Reference = typically-developing children. c Due to an insignificant time effect, the model including
predictors was not run. d Child self-reported well-being was not assessed at T1. e effect size: small < 0.01, medium > 0.09, large > 0.2539.

**Fig1:**
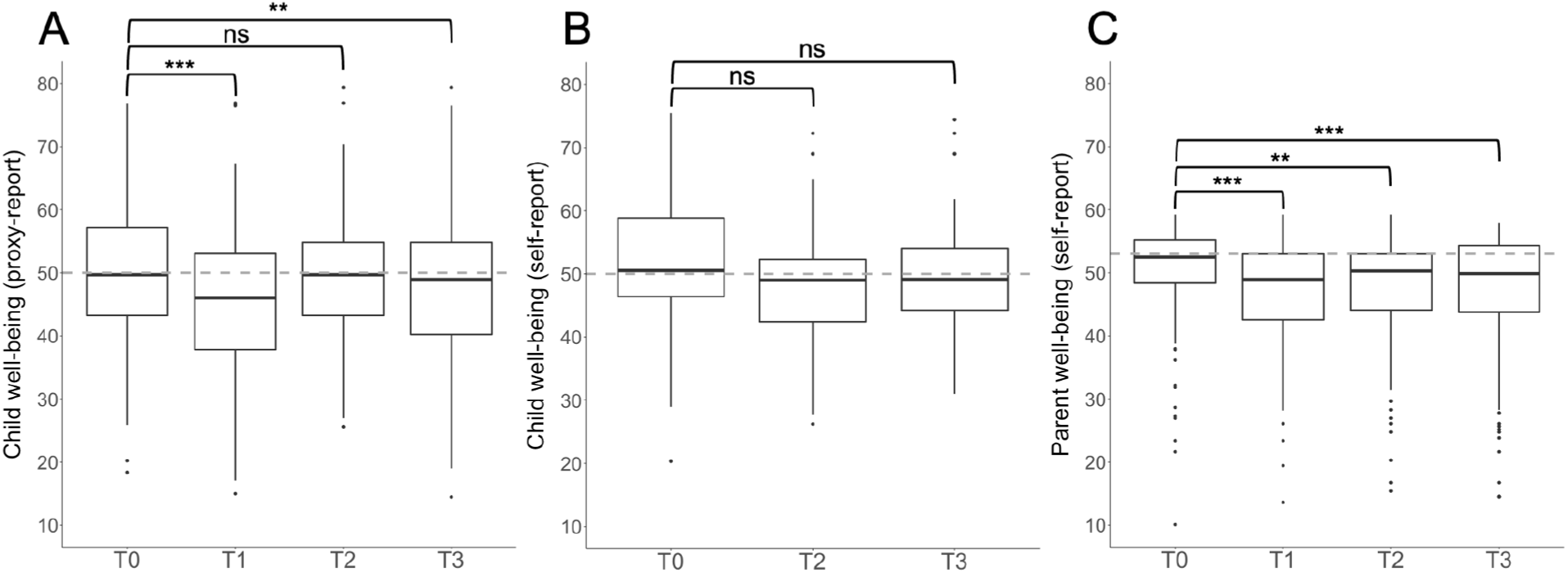
Child proxy-(A) and self-reported (B), and parent (C) well-being before and during the COVID-19 pandemic. *Note*. The box represents the interquartile range. The thick line within the box corresponds to the sample’s median. The grey dashed line represents the normative median (child well-being: *Mdn* = 5033, parent well-being: *Mdn* = 5335). Dots represent outliers. T0 = before the COVID-19 pandemic, T1 = first wave of the COVID-19 pandemic (April–May 2020), T2 = second wave of the COVID-19 pandemic (October– November 2020), T3 = third wave of the COVID-19 pandemic (April–May 2021). Child self-reported well-being was not assessed at T1. Well-being is expressed as T-values. ns = not significant, ^**^ P < 0.01, ^***^*P* < 0.001.

For child self-reported well-being, there was no significant change in well-being across time with small effect size (*R*^*2*^_*B*_(CI-95)*=*0.017(0.007 to 0.051), Fig1B). Self-report and proxy-report correlated weakly at T0 (Spearman’s *r*=0.15, *P*=0.050), T2 (Spearman’s *r*=0.11, *P*=0.343), and T3 (Spearman’s *r*=0.354, *P*=0.001).

Compared to before the pandemic, parent well-being dropped significantly during the first wave of the pandemic and remained significantly lower during the second and third waves (Fig1C). The model’s effect size was small (*R*^*2*^_*B*_(CI-95)*=*0.048(0.028 to 0.087)). Adding the predictors increased the model’s effect size to medium (*R*^*2*^_*B*_(CI-95) *=* 0.132(0.101 to 0.186)). The well-being of parents of children born either VPT or with a CHD was higher than the well-being of parents of typically-developing children. Well-being was independent of parent age. Sparse social support before the pandemic and poor family functioning during the first wave significantly predicted lower well-being. There was no significant interaction between assessment time and group (*P*=0.116), assessment time and social support (*P*=0.350), or assessment time and family functioning (*P*=0.137) on well-being.

## 4. Discussion

This study is the first to investigate the mental sequelae for families one year into the COVID-19 pandemic. The findings provide longitudinal evidence that the well-being of both school-aged children and their parents have been compromised over the first year of the pandemic. Parents of typically-developing children experienced lower well-being than parents of children with complex medical histories before and during the pandemic. Families with sparse social support and poor family functioning are particularly at risk for poor well-being.

Concerns about the mental sequelae of the pandemic for families have been raised since measures were initially implemented to reduce the spread of COVID-19 in spring 2020 [3-7]. However, despite early calls for longitudinal research [7, 41], to date very little is known about the potential long-term mental sequelae for families. Two studies have reported impaired child and parent well-being a few months after the initial wave, suggesting a prolonged negative impact [42, 43]. The current study is the first to link child and parent well-being before the outbreak of the pandemic in March 2020 to the well-being of the same individuals at three time-points over the following year. Tracking these families as the pandemic evolved revealed compromised well-being in both children and parents, with up to one in three experiencing substantially compromised well-being during the first year of the COVID-19 crisis. In addition to confirming the drop in child and parent well-being during the first wave of the pandemic reported by previous studies [8-27], the current study adds an important long-term perspective: Parents reported that the well-being of their children was lower during the first and third waves of the pandemic but not the second. Parents reported that their own well-being dropped during the first wave and remained low during the second and third waves. In Switzerland, schools were open during the second wave, in contrast to mandatory home-schooling during the first wave. Potentially, this contributed to the intermediate recovery of well-being in children and adolescents. However, social distancing and home-office orders were reinforced as the second wave surged. This likely strained parents and continued to compromise their well-being. Children and adolescents themselves reported levels of well-being during the pandemic similar to those before. This is in line with an observed discrepancy between parent and child accounts of child well-being [44]. Importantly, although well-being was preserved at group-level one year into the crisis, one in five children and adolescents self-reported substantially compromised well-being; this is below one standard deviation from the norm. Similarly, a study in adolescents previously found no change in well-being at group-level during the first wave of the pandemic but reported considerable variability between individuals [45].

Consequently, future research investigating subgroups of individuals with different trajectories of well-being over the course of the pandemic and identifying potential predictors of these trajectories will significantly advance understanding of the long-term mental sequelae of this crisis. This should include investigating the positive effects of the pandemic reported by many studies, such as increased family time and reduced stress from fewer obligations related to school or other activities [e.g., 46].

Interestingly, both before and during the pandemic, parents of children with complex medical histories, namely if they were born either very preterm or with a complex congenital heart disease experienced higher level of well-being than parents of typically-developing children. A change in the perception of well-being by adapting internal standards and values, termed a response shift, has previously been argued to underlie above-norm well-being in clinical populations [47, 48]. Important in the context of the pandemic, in the current study, parents of children both with and without complex medical histories experienced similar drops in well-being over the pandemic. Furthermore, families with a member who was at increased risk for a severe disease course in case of infection with Sars-Cov-2 were affected similarly to families without in their well-being during the pandemic. Importantly, individuals with pre-existing medical conditions have, despite retaining their levels of well-being, previously been shown to experience specific concerns related to the ongoing pandemic, including increased fear of contracting the virus, and may thus require specific attention from their health care providers [e.g., 49, 50].

Families with sparse social support and poor family functioning during the first wave of the pandemic were found to be at particular risk of poor well-being. Social factors have previously been identified as contributors to poor well-being during the pandemic [1]. However, these factors are likely not unique to the COVID-19 pandemic because they have been shown to contribute to poor well-being of children and parents in general [e.g, 51, 52]. Even so, the confirmation of social risk factors for poor well-being during the ongoing pandemic is important for identifying families who are at particular risk for long-term mental sequelae. Moreover, social support may be provided not only by the individuals’ social network but also by professionals, including social workers. Thus, strengthening services during and in the aftermath of the current pandemic may prevent long-term mental health sequelae for those at risk.

### 4.1. Limitations

The current longitudinal investigation draws on two cohort studies not originally designed to investigate the mental sequelae of COVID-19 for families. Thus, some limitations require consideration: The questionnaires for the assessments during the pandemic were selected from the study protocol of the cohort studies to allow changes to be investigated. These questionnaires assess well-being rather than mental health symptoms; thus, the conclusions that can be drawn about the prevalence of mental health problems during the pandemic are limited. The study sample includes children with and without complex medical histories and is not representative of the general population of children and parents in Switzerland. However, the longitudinal findings presented here are in line with and complement findings from cross-sectional studies conducted during the initial wave of the pandemic with nationally representative samples that report child and parent well-being compromised in comparison to normative data [11, 25].

The sample size of the current study was relatively small compared to previous cross-sectional studies [e.g., 15, 16, 20]. However, its longitudinal design ensured well-powered analyses owing to within-subject correlations [53].

Participating families come from high and rather homogenous socio-economic backgrounds, as is often seen in prospective cohort studies [54]. This may explain the absence of any effect of SES on well-being, which had been expected from previous findings [1].

Finally, self-reported well-being of children and adolescents was only assessed during the second and third waves of the pandemic but not the first. Thus, no conclusion can be drawn from this study about any potential immediate effects on child self-reported well-being.

### 4.2. Conclusions

This study provides evidence of the long-term mental sequelae of the COVID-19 pandemic for children and their parents, with up to one third reporting substantially compromised well-being one year into the crisis. Even small psychological impacts of the pandemic have been argued to require careful attention as they may pose a substantial public health problem if reproduced across the whole population [2]. Consequently, it is crucial to provide psychological support to those in need alongside the comprehensive economic measures that have been implemented by governments to recover from the ongoing COVID-19 pandemic.

## Data Availability

For re-analyzes of the data set (for different purposes), additional ethical approval (on an individual user and purpose basis) will be required. The authors are happy to support additional ethical approval applications from researchers for access to this data set.

## Declarations

### Code availability

R Code is available from the corresponding author upon reasonable request.

### Authors contribution

ME and FW conceptualized and designed the COVID survey and carried out the analyses and interpretation of the data and drafted and revised the manuscript.

BL and CH conceptualized and designed the original prospective cohort studies and critically revised the manuscript.

OK and ML contributed substantial intellectual content to the interpretation of the results and critically revised the manuscript.

### Ethics approval

The study was approved by the local ethics committee. The procedures used in this study adhere to the tenets of the Declaration of Helsinki.

### Informed consent

All parents gave written informed consent.

### Consent for publication

not applicable.

## Acknowledgement

We thank the EpoKids and the ReachOut study teams for sharing the pre-Covid data, Aziz Chaouch for his statistical advice, Simon Milligan for language editing the manuscript, Minna Törmänen for her input to the Covid survey, and Selina Bürgler, Lesley Ramseier, and Corina Wettach for their support organizing the Covid survey. We thank all parents, children, and adolescents for their continuous support and participation in our studies.

