## Supplementary Fig1 for "Mental sequelae of the Covid-19 pandemic: Well-being one year into the crisis in children with and without complex medical histories and their parents"

**Journal**: European Child and Adolescent Psychiatry

**Authors:** Melanie Ehrler MSc[1,2,3], Cornelia F. Hagmann MD PhD[2,3,4], Oliver Kretschmar MD[2,3,5], Markus A. Landolt PhD[2,3,6,7], Beatrice Latal MD MPH[1,2,3], Flavia M. Wehrle PhD[1,2,3,4]

**Affiliations** [1]Child Development Center, University Children's Hospital Zurich, ZH, Switzerland; [2]Children's Research Center, University Children's Hospital Zurich, ZH, Switzerland; [3]University of Zurich, Switzerland; [4]Department of Neonatology and Intensive Care, University Children's Hospital Zurich, ZH, Switzerland; [5]Department of Pediatric Cardiology, University Children's Hospital Zurich, Switzerland; [6]Department of Psychosomatics and Psychiatry, University Children's Hospital; [7]Division of Child and Adolescent Psychology, Department of Psychology, University of Zurich, Switzerland

**Supplementary Fig1:** Flow chart

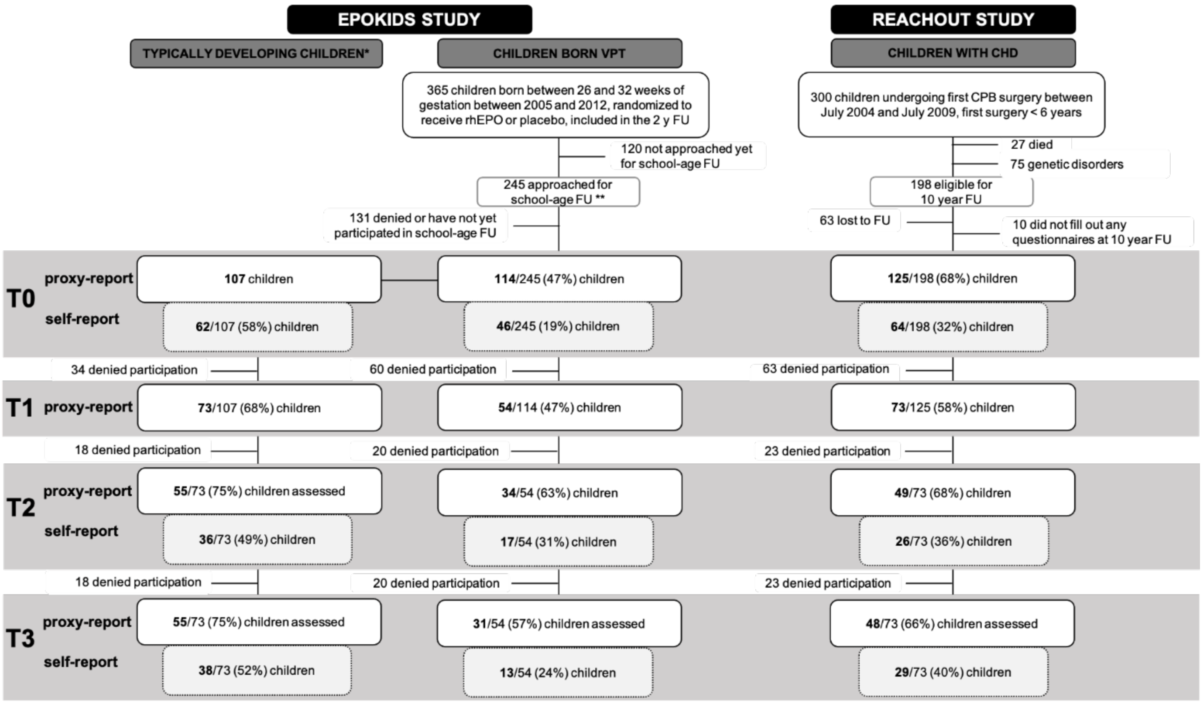


*Note*: Flow chart refers to the child self- and proxy-reports. Numbers of participating parents is lower because some parents reported on more than one child (e.g., T1: 25 parents reported on more than one child, thus, self-reports of 175 parents of 200 children are available). T0 = before

the COVID-19 pandemic, T1 = first wave of the COVID-19 pandemic (April–May 2020), T2 = second wave of the COVID-19 pandemic (October–November 2020), T3 = third wave of the COVID-19 pandemic (April–May 2021).* Typically developing children were recruited as control group for the EpoKids study at school-age **Ongoing study. FU = follow up, CHD=congenital heart disease, CPB = cardiopulmonary bypass surgery. VPT: very preterm. Epokids study1. Reachout study2.


**Supplementary Table 1:** Overview of nationwide measures to reduce the spread of COVID-19 in Switzerland.

| Wave | Nationwide restrictions |
|---|---|
| 1st wave<br>(April/May 2020) | School closure<br>Prohibition of public events<br>Prohibition of private events<br>Closure of nonessential and retail services<br>Reduction of public transport service |
| 2$^{nd}$ wave<br>(October/November 2020) | School closure for adult education only<br>Mandatory face masks in public space for people at the age of 12 years or older<br>Prohibition of public events with >50 people<br>Prohibition of private events with >10 people<br>Prohibition of leisure activities with >15 people<br>Restriction in restaurants to 4 people per table and earlier closing times |
| 3$^{rd}$ wave<br>(April/May 2021) | Prohibition of adult education with > 50 people<br>Mandatory face masks in public space for people at the age of 12 years or older<br>Prohibition of public events with >50 people (inside) and >100 people (outside)<br>Prohibition of private events with >15 people |

| | |
|---|---|
| | Prohibition of leisure activities with >15 people |
| | Inner area of restaurants closed; outdoor area restricted to 4 people per table |

*Note*. Some cantons may have ordered stricter measures. Source: Federal Office of Public Health (FOPH) Switzerland https://www.bag.admin.ch/bag/en/home/krankheiten/ausbrueche-epidemien-pandemien/aktuelle-ausbrueche-epidemien/novel-cov/massnahmen-des-bundes.html (accessed July 5th 2021)
